# Global and regional associations of enteric infections and asthma

**DOI:** 10.1101/2024.05.21.24307671

**Authors:** Duan Ni, Ralph Nanan

## Abstract

Asthma is one of the most common chronic diseases globally. There are various risk factors for asthma, including respiratory infections, like rhinovirus and respiratory syncytial virus infections. Nevertheless, impacts from non-respiratory tract infections on asthma remain unresolved. Globally, enteric infections are the most common infections. Their target organ, the intestinal system, shares the same embryonic origin, the endoderm, as the respiratory tract. These led us to inspect the potential associations between enteric infections and asthma.

We analyzed three independent epidemiological datasets with the cutting-edge generalized additive model (GAM). Consistently, in two regional datasets (America and Japan) and a global dataset, we found that enteric infection incidence was generally linked to increased asthma disease burden.

This is the first study to our knowledge to comprehensively elucidate the positive associations between enteric infections and asthma. Our findings would be instructive to both clinical practice and mechanistic study for asthma.

---

Asthma is one of the most common chronic diseases worldwide, with considerable disease burden in developed countries [1, 2]. Established risk factors for asthma other than genetics include asthmagen exposure, obesity, smoking and air pollution. Socioeconomic factors are critically implicated as well [1, 3]. Additionally, respiratory tract infections, triggered by rhinovirus and respiratory syncytial virus (RSV), have been linked to asthma [2]. However, the influences of non-respiratory tract infections on asthma remain elusive. Globally, gastrointestinal infections are the most prevalent, even exceeding respiratory tract infections [4]. Given their significant disease burden and the shared endodermal origins of their target organs [5], namely, the respiratory and gastrointestinal systems, we aimed to inspect the potential associations between enteric infections and asthma.

For this purpose, we interrogated the relationship between the disease burden of enteric infections and asthma, harnessing the unprecedented comprehensive epidemiological data from the Global Burden of Disease (GBD) database (https://www.healthdata.org/research-analysis/gbd). Age-standardized incidence rates for enteric infection (EI%) and asthma (asthma%) for both sexes from countries and areas under study were first collated. They covered over 150 countries from 1990-2018. Annual gross domestic product (GDP) per capita was used as reflections of socioeconomic status. Global GDP data was obtained from the Maddison project [6]. GDP data for individual states in America were from the U.S. Department of Commerce, Bureau of Economic Analysis and GDP data for individual prefecture in Japan were from the Organisation for Economic Co-operation and Development (OECD). EI% and asthma% were first log10-transformed and then modelled with generalized additive model (GAM) with the *mgcv* package [7], a powerful analytical tool to systematically elucidate the effects from multiple parameters and their interactions, including non-linear effects. GDP was adjusted as a confounder, while the regions from which the data originated were accounted as random effects. For most analyses, the model below is favored based on Akaike information criterion [8], in which the main effects from EI%, GDP and time are modelled with smooth term s() and their interactions are modelled with tensor production interaction ti(). This model revealed the interactive effects among EI%, GDP and time on asthma%.

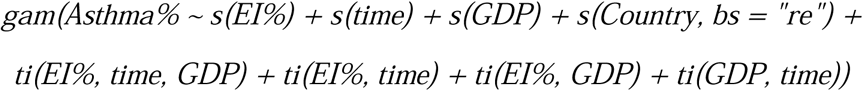

While for the modelling for Japan, the time factor was excluded, considering that data were only available at two timepoints.

We first focused on individual countries with high-resolution regional data. EI% and asthma% from 50 states in America from 2008 to 2017 were analyzed. As shown in Figure 1A, EI% strikingly correlated with increased asthma%, independent of GDP. Analysis of a separate independent dataset across 47 prefectures in Japan at timepoints of 1999 and 2018 yielded similar results (Figure 1A).

**Figure 1.**
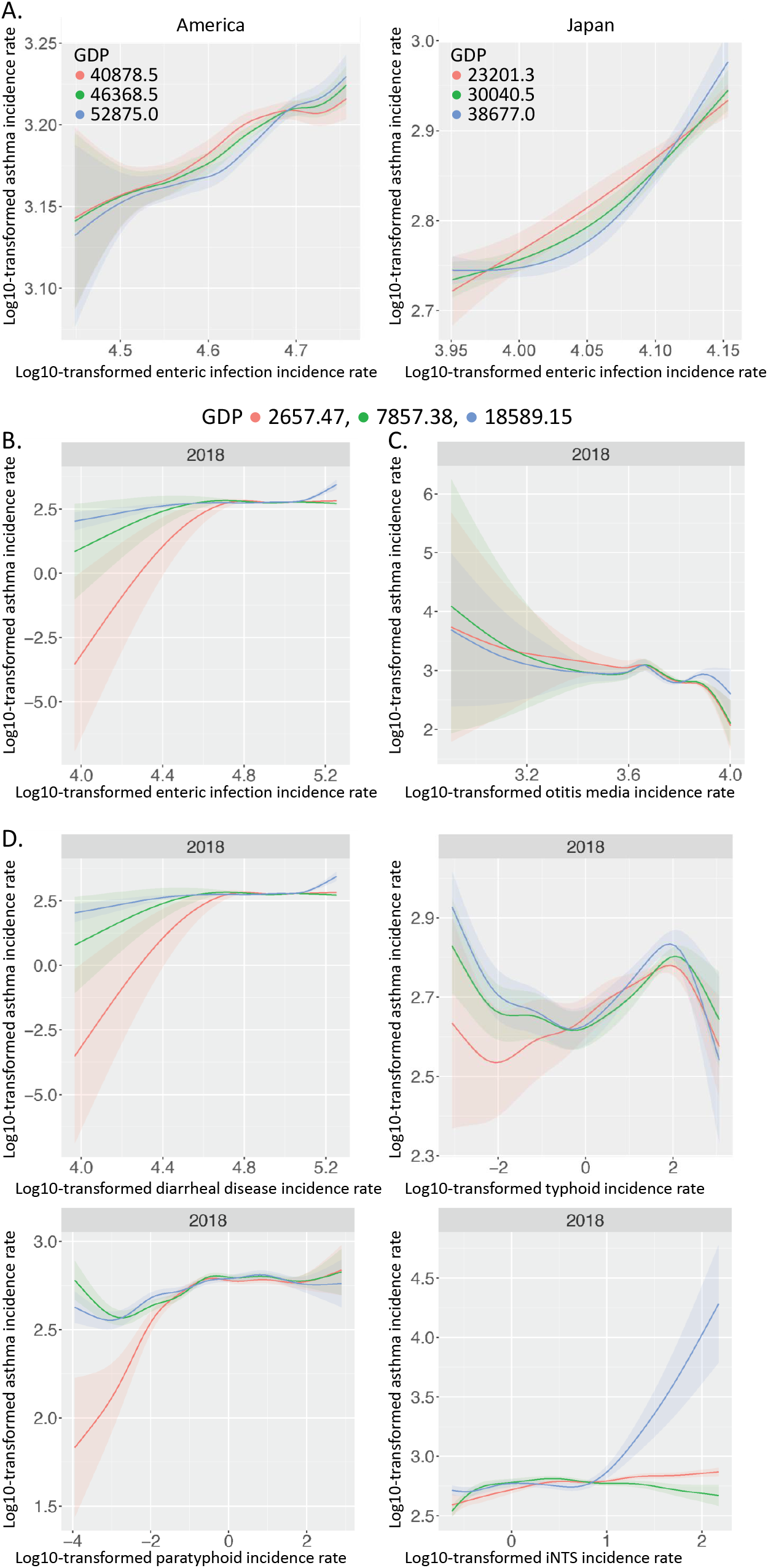
**A**. Modelled effects of log10-transformed enteric infection incidence rate (EI%) on log10-transformed asthma incidence rate (asthma%) across 50 states in America (left) and 47 prefectures in Japan (right), stratified by 25% (red), 50% (green) and 75% (blue) quantiles of GDP. **B**. Modelled effects of EI% on asthma% globally, stratified by 25% (red), 50% (green) and 75% (blue) quantiles of GDP. **C**. Modelled effects of log10-transformed otitis media incidence rate on asthma% globally. **D**. Modelled effects of log10-transformed incidence rates of diarrheal disease, typhoid, and paratyphoid and invasive non-typhoidal Salmonella (iNTS) infection on asthma% globally.

We next approached the analyses at a global scale and analyzed GBD data for asthma and enteric infections spanning over 150 countries from 1990 to 2018. Figure 1B showed the modelling for 2018 as the most recent representative results, with GDP stratified at 25% (red), 50% (green) and 75% (blue) quantiles. In alignment with aforementioned results, EI% generally correlated with elevated asthma%, most pronounced in low GDP countries (red line) (Figure 1B). Interestingly, such correlations seemed to reach plateaus at about log10-transformed EI% of 4.6 (i.e. EI%∼39810), but slightly increased again in high GDP countries (blue line) beyond 5.1 log10-transformed EI% (i.e. EI%∼125892).

In comparison, a common respiratory tract infection, otitis media, only mildly correlated with reduction in asthma% (Figure 1C).

There are 4 major types of enteric infections documented in the GBD dataset (diarrheal diseases, typhoid, paratyphoid and invasive non-typhoidal Salmonella (iNTS) infection). Generally, with results from 2018 shown as representatives in Figure 1D, diarrheal diseases seemed to be the main contributor to the positive associations between EI% and asthma%, while typhoid, though previously reported as a triggering factor for asthma [9], was not strongly correlated. Intriguingly, in low GDP countries (red line), paratyphoid was also associated with asthma% elevation, while in high GDP countries (blue line) iNTS infection was linked to higher asthma%.

Collectively, here, we present to our knowledge the first comprehensive analyses of the associations of various enteric infections and asthma. At both global and regional levels, we delineated the positive correlations between EI% and asthma%, independent of GDP and time.

Some previous studies documented the links between asthma with intestinal manifestations [10], though their causality remains unclear. Developmentally, respiratory and gastroenteric tracts are from the same embryonic origin, endoderm. Whether there might be some shared endodermal intrinsic defect that could account for the correlations between these pathologies requires more in-depth investigation. In contrast, little correlation was found for otitis media and asthma, possibly due to the complex developmental origins of the middle ear, forming from all three embryonic germ layers [11].

Another possible explanation for the connection between intestinal infection and asthma might be via the gut microbiome. It is well-established that gut dysbiosis is linked to asthma development [12]. Infection of the enteric system inevitably disrupts gut homeostasis and microbiome, which might contribute to the observed increase in asthma%.

Our present analyses only cover 4 main types of enteric infections, diarrheal diseases, typhoid, paratyphoid, and iNTS. In contrast to these diseases which show no clear bias towards a Th2 phenotype [13, 14], critical for asthma pathogenesis, some intestinal helminth infections, like gastric helminthiasis in children, were positively associated with asthma [15], possibly because of the Th2 responses mounted upon infections. Hence, more detailed interrogations regarding the types of infectious agent and the severity of enteric infections are needed to decipher the potential underlying mechanistic links.

In conclusion, we identified in three independent epidemiological datasets on global and regional levels associations between enteric infections and asthma. This warrants more surveillance of their concurrence in clinical practice and could prompt more in-depth mechanistic discoveries towards their immunopathology.

## Data Availability

All data produced in the present study are available upon reasonable request to the authors

## Ethics statement

Not applicable

## Conflicts of interest

The authors declare no conflicts of interest.

## Support statement

This study was supported by the Norman Ernest Bequest Fund.

## Notes

### Competing Interest Statement

The authors have declared no competing interest.

